# Associations of the BNT162b2 COVID-19 vaccine effectiveness with patient age and comorbidities

**DOI:** 10.1101/2021.03.16.21253686

**Authors:** Idan Yelin, Rachel Katz, Esma Herzel, Tamar Berman-Zilberstein, Amir Ben-Tov, Jacob Kuint, Sivan Gazit, Tal Patalon, Gabriel Chodick, Roy Kishony

## Abstract

Vaccinations are considered the major tool to curb the current SARS-CoV-2 pandemic. A randomized placebo-controlled trial of the Pfizer BNT162b2 vaccine has demonstrated a 95% efficacy in preventing COVID-19 disease. These results are now corroborated with statistical analyses of real-world vaccination rollouts, but resolving vaccine effectiveness across demographic groups and its interaction with comorbidities is challenging. Here, applying a multivariable logistic regression analysis approach to a large patient-level dataset, including SARS-CoV-2 tests, vaccine inoculations and personalized demographics, we model vaccine effectiveness at daily resolution and its interaction with sex, age and comorbidities. Vaccine effectiveness gradually increased post day 12 of inoculation, then plateaued, around 35 days, reaching 95.0% [CI 93.4%-96.3%] for all infections and 99.5% [CI 97.0%-99.9%] for symptomatic infections. While effectiveness was on average uniform for men and women, it declined mildly but significantly with age especially for males. Effectiveness further declined for people with type 2 diabetes, COPD, and immunosuppression, as well as cardiac disease in females. Quantifying real-world vaccine effectiveness, including both biological and behavioral effects, our analysis provides initial measurement of vaccine effectiveness across demographic groups.

---

The mRNA-based BNT162b2 COVID-19 vaccine (‘Pfizer vaccine’) has demonstrated a 95% efficacy in preventing COVID-19 in phase III randomized placebo-controlled trial, with early protection from the disease evident already 12 days after the first dose ^1,2^. Despite the benefits of a controlled trial, it is limited in resolution due to restrictions during the recruitment process and the relatively small sample size. For example, the phase III trial did not allow the participation of immunosuppressed patients, or patients with unstable chronic conditions^3^. The rapid national vaccination rollout in Israel provides an opportunity to test the effectiveness of the vaccine in real-world prevention of SARS-CoV-2 infection and disease across a diverse population. However, estimating the real-world effectiveness of the vaccine is challenging due to strong temporal and spatial epidemic patterns, and association of testing with vaccination.

Several approaches have been applied to tackle these challenges. First, using the vaccinated population as both the experiment and control groups - comparing the infection incidence for the vaccinated population starting at the early protection period (starting at day 12) with incidence during the first 11 days after the first dose, when patients are not yet protected - has identified an effectiveness of over 50% in later days^4^. Models accounting for disease dynamics in the general population suggest a 66-85% reduction in infections with over 90% reduction in severe hospitalizations^5^. In a different effort, leveraging known associations of vaccination with population characteristics such as age and geographical location, population-wide associations of infection incidence and hospitalizations with vaccination rates were identified^6^. Finally, a comprehensive comparison of infection and disease incidences between a vaccinated population and a demographically and clinically matched unvaccinated control group has yielded an estimation of effectiveness similar to the randomized-control trial, and showed a reduction in vaccine effectiveness in patients with multiple comorbidities^7^. Yet, quantifying the association of vaccine effectiveness with multiple patient-specific attributes and resolving behavioral versus biological effects, while controlling for patient demographics and the dynamically varying volume of the epidemic, remains challenging. Indeed, vaccinated and unvaccinated patients could also differ in their rate of testing. In recent years, machine-learning based approaches have become powerful in quantifying vaccine effectiveness in reducing infections per-test while accounting for confounding variables^8–11^. Here, generalizing these approaches, we build a multivariable logistic regression analysis for both per-day and per-test infections, that allows us to calculate infection risk for different post-vaccination time ranges, while adjusting for spatial and temporal patterns of the epidemic and for patient-specific characteristics such as age, sex and comorbidities. Adding interaction terms of time from vaccination with sex, age and comorbidities, we further resolve the associaitons of these patient attributes with vaccine effectiveness. Applying this methodology on infections observed either per-day or per-test, and comparing associations at later times following vaccination with associations observed at early days, before any presumed immunological protection, help us to further resolve behavioral and biological effects of the vaccine.

Between 19th of December 2020 and February 25th 2021, Maccabi Healthcare Services (MHS) has vaccinated more than 1.2 million out of almost 1.8 million of its 16 years old and older population, as part of a national rapid rollout of the vaccine. We collected, for each member of MHS, anonymized data including vaccination dates, results of any oral-nasopharyngeal SARS-CoV-2 tests, age, sex, and city of residence, as well as tagging of comorbidities including: cardiovascular disease, high blood pressure, body mass index (BMI), chronic kidney disease (CKD), chronic obstructive pulmonary disease (COPD), immunosuppression and type 2 diabetes (Methods). We then performed a logistic regression analysis on a matrix where each line (“observation”) corresponds to a given person at a given calendar day (67 calendar days, over 93 million observations after exclusion, Methods). The outcome of each of these observations indicates whether or not the specific person had a positive test on the specific calendar day (0/1). The features include the calendar day, vaccinated versus unvaccinated, the number of days post-vaccination and patient-specific characteristics including sex, age, place of residence, and patient’s comorbidities. Finally, to characterize vaccine effectiveness, we also included interaction terms of age, sex and comorbidities with vaccination for three distinct post-inoculation time periods (1-11, 12-28, 29-50 days; Methods). We consider two models, one that includes all 67 calendar days for each patient (thereby predicting the “per-day” risk of a positive test result) and the other that includes for each patient only the calendar days in which they were tested (thereby predicting the “per-test” risk of a positive test result).

The odds ratio of infection for different days following vaccination, compared to unvaccinated reference, showed a gradual decrease in infection rate starting at day 12, ultimately plateauing, following 35 days post first inoculation, at levels of approximately 95% for the per-test model and 96% for the per-day model. The model coefficients for post-vaccination days provide the odds ratios for infection relative to an unvaccinated reference (Figure 1). In the per-day model, we observed an initial negative association of infections with vaccination at days 1-5, coinciding with a decrease in tests on these initial post-inoculation days (Supplementary Fig. 1a, presumably due to a behavioral tendency to avoid testing immediately following vaccination). Guidelines required a second dose be given 21 days after the first dose. Adherence to guidelines was high with more than 98% of patients administered with the first dose being administered with two doses, most frequently on day 21 (Supplementary Fig. 2). Consistent with the phase III placebo-controlled trial, an initial decrease in infection rate is observed at day 12. These rates further decrease until a plateau is reached following 35 days post-inoculation at levels of 96.0% (CI 94.7%-97.0%) for days 44-50. Vaccinated individuals are not required to get tested for epidemiological reasons (e.g. after contact with a COVID-19 patient). Therefore, effectiveness at later days may be overestimated. To alleviate this potential bias, we used the per-test model, considering the risk of infection per test. We found an initial apparent effectiveness already in days 1-11 (cyan period), explained by a time-independent (existing even prior to vaccination) positive association of the vaccinated group with testing (Supplementary Fig. 1a; likely due to association with access and tendency for treatment). Focusing on later days (magenta and purple periods), vaccine effectiveness gradually increased, reaching a plateau at a similar level of 95.0% (CI: 93.4%-96.3%).

**Figure 1.**
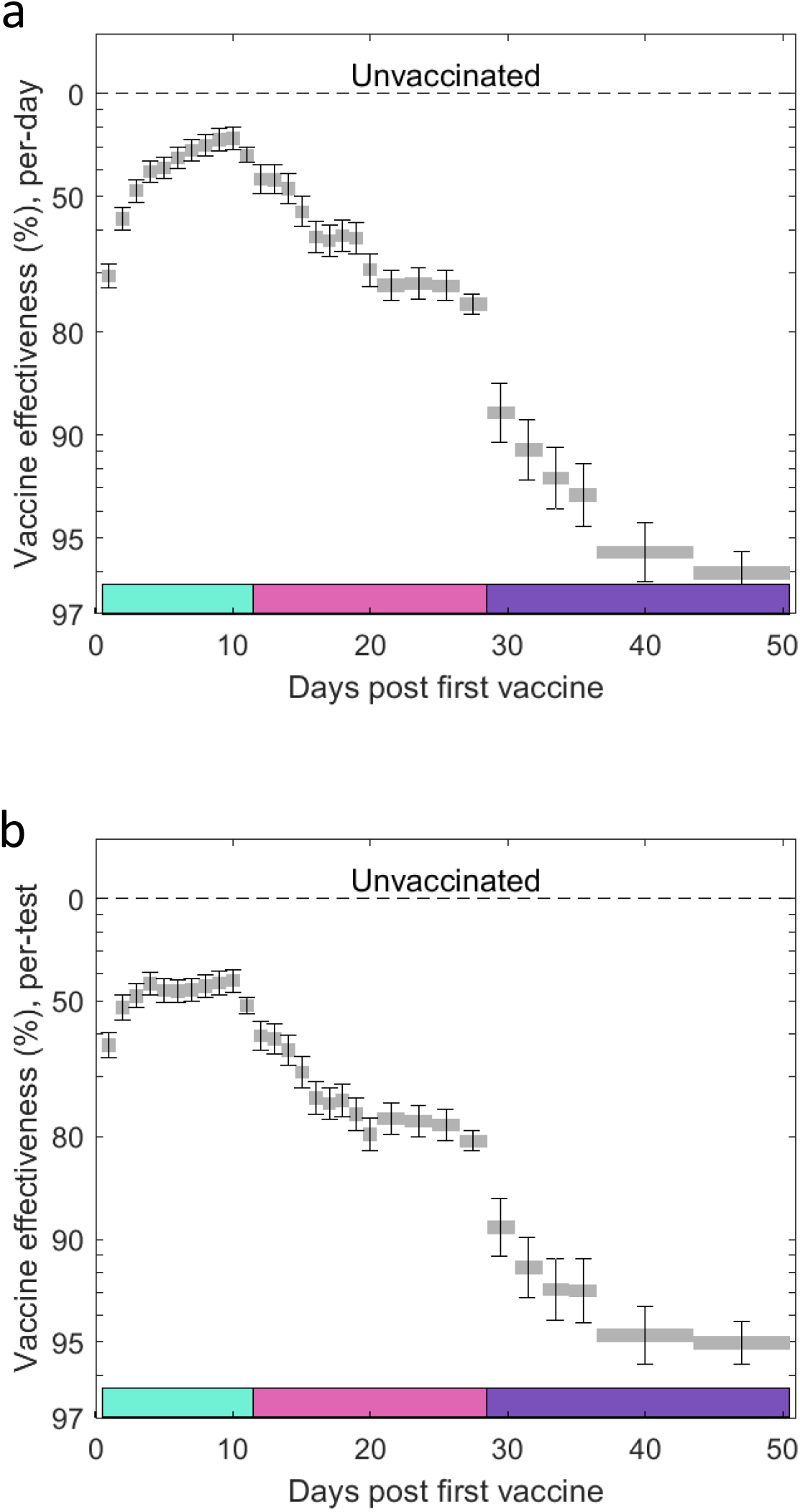
Vaccine effectiveness following vaccination at daily resolution. Vaccine effectiveness (VE, Methods) for per-day (a) and per-test (b) infections, for different days post-vaccination. Error bars indicate one standard error (Methods). Dashed horizontal line at 1 indicates infection risk of the unvaccinated reference group. Three post-vaccination time periods are indicated: days 1-11 (cyan), 12-28 (magenta), 29-50 (purple). Number of cases: n_vaccinated_=1,721,377, n_unvaccinated_= 459,069.

We next asked whether and to what extent vaccination effectiveness might vary across demographic characteristics and whether it may be associated with certain patient comorbidities. We considered the coefficients for interactions of sex, age and comorbidities with 3 distinct post-vaccination time ranges: 12-28 and 29-50 days, as well as with days 1-11 as a pre-immunization control period (where vaccination is not presumed to have a biological effect). In both the per-day and per-test models, the odds ratio of vaccine effectiveness with sex, age, and any of the comorbidities considered was at most two-fold, indicating that the vaccine remains effective across demographics and comorbidities (Fig. 2). For sex, vaccine effectiveness for men and women was highly similar (Odds ratio for male versus female of 1.04 [CI: 0.88-1.22] for per-day and 1.12 [CI: 0.95-1.32] for per-test models). For age, considering the per-day model (incidence rate), vaccine effectiveness seemingly declined at older age (81-90 age group), yet similar associations also appeared for this age group in the pre-immunization period (Fig. 2A, top, cyan), suggesting an underlying behavioral effect (pre-existing tendency for increased testing in vaccinated versus unvaccinated patients was indeed amplified for older patients, Supplementary Fig. 1a). Correcting for this effect, the per-test model diminished much of the interaction of age with vaccine, revealing a mild decrease in vaccine effectiveness with age at the post-immunization periods (days 29-50, Odds ratio 0.74 [CI: 0.52-1.06] for 81-90 years old relative to the reference age group). For comorbidities, considering the per-day model, blood pressure, COPD, immunosuppression and type 2 diabetes reduce vaccine efficacy, yet they also interact, to a lesser extent, with vaccination at the control pre-immunization time period (Fig. 2b, top). These interactions at the pre-immunization period can possibly be explained by the relatively lower tests per day of patients with one or more comorbidities (Supplementary Fig. 1b). Accordingly, in the per-test model, a reduction in vaccination effectiveness is seen uniquely for the later, post-immunization, time periods (Fig. 2b, bottom, days 29-50; Odds ratio 0.73, [CI: 0.59-0.91] for type 2 diabetes; 0.67 [CI: 0.53-0.83] for immunosuppression and 0.55 [CI: 0.38-0.80] for COPD).

**Figure 2.**
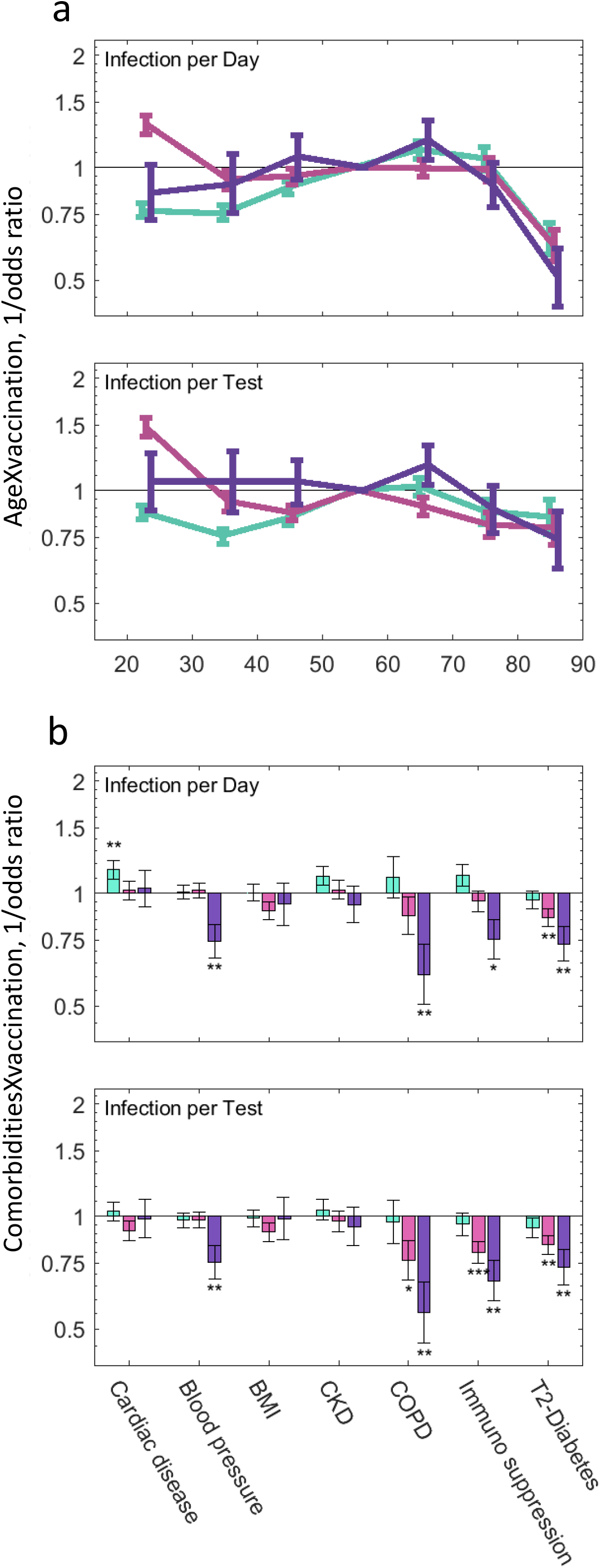
Odds ratio of infection for the interaction of age and comorbidities with vaccination. Odds ratio for per-day (top) and per-test (bottom) infections, for interaction of age (a) and comorbidities (b) with vaccination at three different post-vaccination time periods: 1-11 (cyan), 12-28 (magenta) and 29-50 days (purple). Values higher than 1 (horizontal line) indicate greater vaccine effectiveness, while values lower than 1 indicate reduced effectiveness. Error bars indicate one standard error of the corresponding logistic regression coefficients. * p<0.05, ** p<0.01, *** p<0.001.

The interaction of comorbidities and age with vaccination differed for males and females. Considering the coefficients of interactions of age with each of the 3 post-vaccination time ranges separately for the two sexes, we found that the decrease in vaccine effectiveness observed for 81-90 years old individuals in the per-test model, was only significant for the male population. Males 81-90 years old had an odds ratio of 0.47 [CI:0.28-0.79] (Supplementary Fig. 3). These results are consistent with the overall increased risk and severity of the disease in older males^12^. Conversely, cardiac disease and blood pressure were negatively associated in the per-test model with vaccine effectiveness at the later post-immunization period only among females (0.66 [CI: 0.47-0.94] and 0.73 [CI: 0.54-0.99], respectively) while no significant association was identified among males (1.25 [CI: 0.92-1.72] and 0.77 [CI: o.58-1.03], respectively). These comorbidities are indeed known to manifest differently for females and males^13^.

Vaccine was even more effective in preventing symptomatic infections. A subset of the tests had an associated physician referral indicating symptoms (referrals are not required and were usually not issued; only 2.2% of the tests had an associated referral which also indicated symptoms). We repeated the logistic regressions with the same features as above, but when calculating the odds ratio for a positive test with a symptomatic referral (Fig. 3, Methods). Vaccine protection against symptomatic infections was higher than the effectiveness observed for all infections both when considering risk of infection per day (effectiveness in days 44-50 post-vaccination was 99.6% [CI: 97.5%-99.9%] for symptomatic infections versus 96.0% [CI: 94.7%-97.0%] for all infections, P<0.02) and per test (99.5% [CI: 97.0%-99.9%] versus 95.0% [CI: 93.4%-96.3%], P<0.02, Methods).

**Figure 3:**
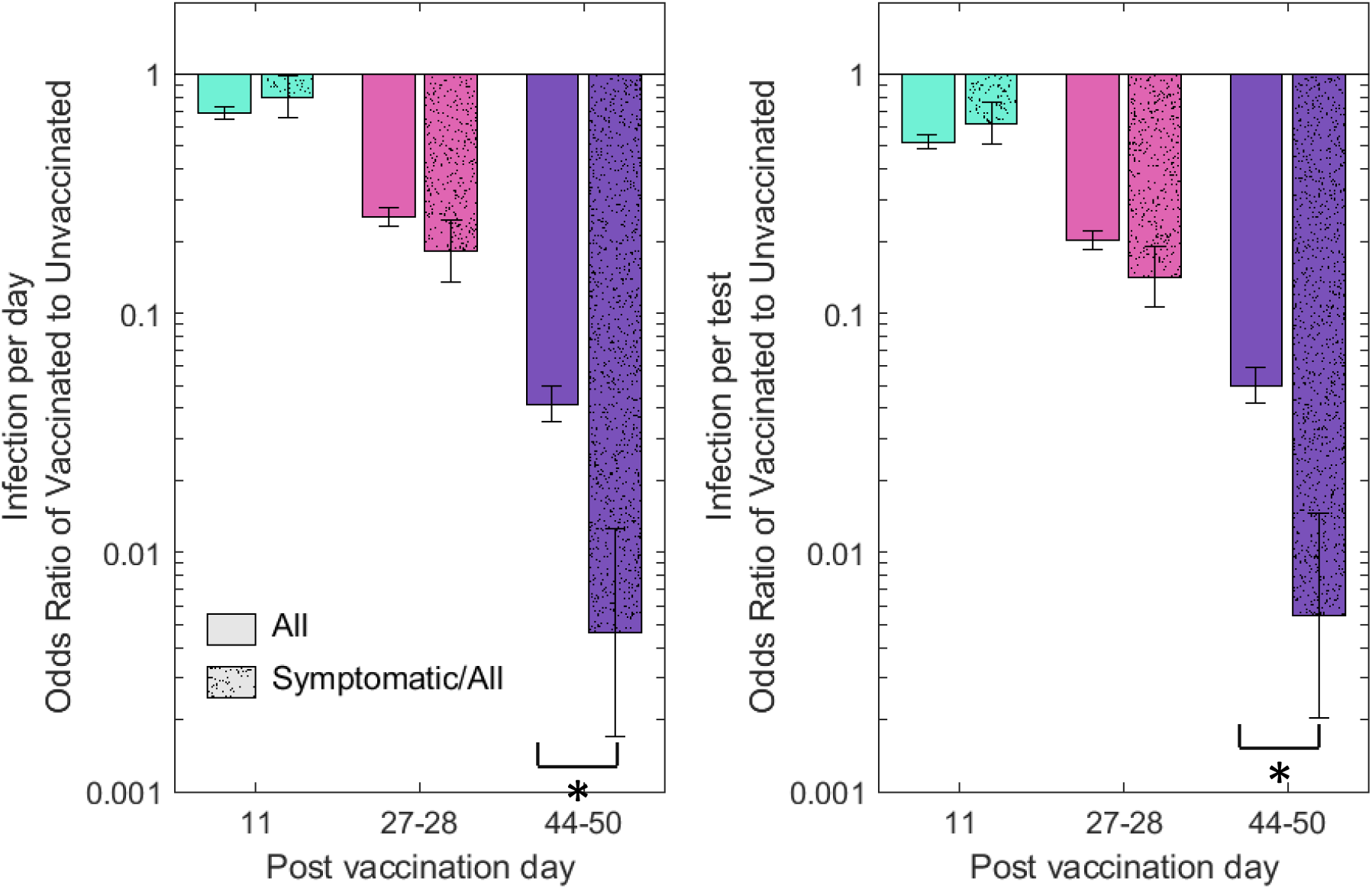
Vaccine effectiveness for symptomatic versus all infections. Vaccine effectiveness (VE, Methods) in a per-day (a) and per-test (b) model of all (solid bars), or symptomatic (hashed bars) infections at days 11 (cyan, before immunity control), 27-28 (magenta) and 44-50 (purple) post first vaccine inoculation.

Our study has several limitations characteristic of observational studies. First, our data reflects uncontrolled non-random testing and non-random vaccination, both potentially biased across the population. Second, the vaccinated population may differ from the unvaccinated population in its general health status, in its risk of being infected and in its health-seeking behavior. These differences may be both inherent, pre-existing even prior to vaccination, and time-dependent as a result of vaccination itself. Third, during the study period, several viral variants were circulating in Israel. Although the vaccine is expected to be potent against B.1.1.7 ^14^, which was the most common one ^15^, it is possible that additional variants introduced biases to our estimations of effectiveness across subpopulations, especially if vaccinated at different phases of the epidemic. Fourth, our analysis of vaccine effectiveness in preventing symptomatic disease may be biased by a larger fraction of mild, and therefore, unreported, symptomatic disease among the vaccinated population.

While behavioral biases listed above are an inherent challenge in any real-world data, our analysis offers ways to detect and partially account for such behavioral differences. Using a model for tests per day (independently of the test results), we find that vaccinated patients are tested more frequently than unvaccinated even well before their first vaccine inoculation, that this effect is substantially increased for older patients, and that this intrinsic tendency for testing strongly declines at the first few days following vaccination. Using the per-test model, we adjust for differences between the populations in their tendency to get tested ^10^. Such correction removes pre-existing biases, though it may also over-correct as decision to get tested is driven not only by behavioral tendencies but also by the infection itself. The per-test removal of behavioral effects is further corroborated by contrasting the associations identified for the late immunization period (after day 28) with any possible associations in the pre-immunization period where biological effects are not expected. We note also that potential behavioural biases are somewhat minimized in our dataset due to the rapid pace of freely offered vaccination, together with freely offered laboratory testing for HMS members. The high disease rate during the study period also makes this dataset suited for analysis, especially as we consider calendric dates to account for the dynamics of the epidemic wave.

Our analysis of the Pfizer BNT162b2 vaccination and infection records identifies an onset of infection prevention effectiveness at 12 days after the first-inoculation in a two-dose vaccine regimen, gradually increasing till a plateau at 95.0% [CI 93.4%-96.3%] for all infections and 99.5% [CI 97.0%-99.9%] for symptomatic infections. While the effectiveness against symptomatic infections is slightly higher than the efficacy reported in the clinical trial, providing daily time resolution, our analysis reveals that these high levels of vaccine effectiveness are only fully reached following day 35, in agreement with the expected period of two weeks after the second dose^16^. Comparing effectiveness across age groups, we find that although vaccine effectiveness is relatively similar for age groups between 16-80 years old, slightly lower effectiveness is observed for older patients (81-90 years old). While on average vaccine effectiveness was almost the same for males and females, separately analysing females and males, we find that the age related discrepancy can be largely attributed to males, where effectiveness decreases for the elderly. We also find that specific chronic comorbidities, including high blood pressure, COPD, immunosuppression and type 2 diabetes, are negatively associated with vaccine effectiveness. These results add to previous reports regarding lower vaccine effectiveness for diabetic patients ^4^ and patients with multiple coexisting conditions ^7^. Importantly, our approach allows quantifying these associations, even for less frequent comorbidities, such as for immunosuppressed patients, who were not represented in the clinical trial^1^. Furthermore, our analysis shows sex-specific interactions with certain comorbidities, mosty notably showing reduced effectiveness specifically for women with cardiac disease or high blood pressure. We emphasize that while these attenuations are significant, even for people with attenuating factors the vaccine is still highly effective. Our methodology provides a unified framework for analyzing vaccine effectiveness and its dependence on patient’s attributes from dynamic spatially distributed datasets and may help quantify the effectiveness of other vaccines and how it may vary in longer time scales.

## Methods

### Data collection

Anonymized electronic health records were retrieved for the study period December 1st 2020 - February 25th 2021 for MHS members older than 16 (1.79 million). These records include: (a) Patient demographics, indicating for each MHS member: a random ID used to link records, year of birth, sex, coded geographical location of residence at neighborhood resolution, and chronic comorbidities including: cardiovascular disease, type 2 diabetes, high blood pressure, immunosuppression, high BMI, chronic kidney disease (CKD) and chronic obstructive pulmonary disease (COPD). (b) Test results, indicating for any SARS-CoV-2 RT-qPCR test performed for MHS members: the patient random ID, the sample date, and an indication of positive and negative results (total 1,163,799 tests, with 66,944 positive results). (c) Referral for SARS-CoV-2 test, indicating the patient random ID, the referral date and a reason for referral (83,714 referrals, 26,095 of which indicated symptomatic infection; most tests are performed without a referral). (d) Vaccination, indicating patient by random ID and their dates of first and second inoculations with the BNT162b2 mRNA COVID-19 (1.27 million vaccinees). While vaccination rollout started on December 19th 2020, test records were obtained for the period starting on December 1st to establish a baseline of tests per day. We excluded from the study patients who had any one of the following indications: (a) age above 90 (excluding 9,792 from the total of 1,799,240 individuals); (b) a positive test prior to the study period (excluding an additional 65,939 individuals).

### Per-day multivariable logistic regression model for infection

We performed a logistic regression for the risk of a positive test for each patient in each calendar day (up to their first positive test if any). Given 1.79 million patients, and 67 calendar dates (Dec 20 - Feb 25), we have >120 million patient-cross-date observations (PxD). For each PxD, the predicted variable is Y=0/1, indicating whether the specific patient had a positive result on the specific calendar date (1 - positive test, 0 - no test or a negative test). The features for each PxD observation (the “X” matrix) include: (a) Sex (0/1, Female/Male), (b) Age (length-6 dummy-variable vector designating 0/1 for age bins: 16-30, 31-40, [41-50], 51-60, 61-70, 71-80 and 81-90, with the [41-50] variable omitted such that all-zeros indicates this age bin as the reference), (c) comorbidities (length-6 binary vector indicating absence/presence of each comorbidity), (d) calendar date and neighborhood of residence (target encoding) and (e) post-vaccination time, encoded as post-vaccination period (PVP), and post-vaccination day (PVD). The PVP is a length-3 dummy-variable vector designating 0/1 for periods 1-11, 12-28, 29-50 days post first-dose inoculation (corresponding to the cyan, magenta, purple periods in Figures 1–3), with [0,0,0] indicating unvaccinated patients or patients at dates prior to their first-dose inoculation (we specifically omit PxD corresponding to days beyond the 50 days limit or to the 2 weeks prior to vaccination where behavioral effects are suspected, Supplementary Fig. 1). The PVD is a dummy-variable vector representing post-inoculation days 1, 2, 3, 4, 5, 6, 7, 8, 9, 10, [11], 12, 13, 14, 15, 16, 17, 18, 19, 20, 21, 22, 23-24, 25-26, [27-28], 29-30, 31-32, 33-34, 35-36, 37-43, [44-50]. To avoid over-parameterization due to dependency among PVD and PVP, the last bins of PVD in each of the three periods (bins [11], [27-28], and [44-50]) are omitted, such that PVD is a length-28 binary vector and all-zero represents the first day of the period indicated by PVP (or unvaccinated if PVP is also all-zeros). This coding of post-vaccination day into period and days within the period allows quantifying interactions of Sex, Age and Comorbidities with vaccination, by including the interaction terms: Sex*PVP (length-3 binary vector), Age*PVP (length-18 binary vector), and Comorbidities*PVP (length-18 binary vector). Together, the model includes 83 features. In practice, to allow faster computation and to fit the data within a reasonable computer memory, only unique lines of this matrix are stored and, for each such unique line, the total number of PxD’s (or PxD’s with a test) and total positive result are calculated. The model is then solved in Matlab with the glmfit function.

### Per-test multivariable logistic regression model for infection

Starting with the feature matrix and outcome described above, we omitted all PxD lines at which a test was not performed (namely keeping for each patient only observations on dates in which they were tested). We then performed the logistic regression as above on this trimmed input matrix. The model thereby calculates the probability for a positive result given that a patient had performed a test on a given date. For symptomatic infections the same feature matrix was used (same PxD lines), but the dependent variable indicated a positive test with a symptomatic referral.

### Regression model for rate of testing

We repeated the logistic regression with the full PxD input matrix, with the following changes: (a) the outcome Y for each PxD specifying whether or not a test was performed for the specific patient at the specific date (0/1); (b) adding PVD bins corresponding to pre-vaccination times as early as 27 days prior to vaccination, as indicated in Supplementary Fig. 1. (c) excluding PxD of patients at days prior to the most negative time bin [(−26)-(−27)], such that the unvaccinated control group includes only people who did not get vaccinated at any point during the study period; (d) ran the model separately on each age group. The results of this model, predicting the probability of testing rather than the test result, are presented in Supplementary Figure 1.

### Model interpretation

Odds ratio for infection following vaccination (Fig. 1) were calculated as 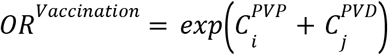, where 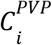 and 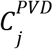 are the regression coefficients of the model for the post-vaccination period *i* and post-vaccination day *j* (except substituting 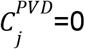 for *j* corresponding to the reference post-vaccination days 11, 27-28, or 44-50). For example, the odds ratio of infection in day 38 post-vaccination is 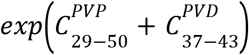, and for day 27, which is in bin 27-28 which is a reference bin for period 12-28, the odds ratio is 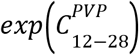. These odds ratios for infection were transformed into vaccine effectiveness (VE), VE=1-OR^*Vaccination*^. The effect of comorbidities on vaccine effectiveness was calculated as 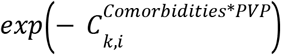, where *k* designates one of the 6 comorbidities and *i* designates one of the 3 post-vaccination periods (cyan, magenta, purple; Fig. 2, bottom). Similarly, the effect of Age on vaccine effectiveness was calculated as 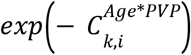, where k designates one of the 6 age bins and *i* designates one of the 3 post-vaccination periods (cyan, magenta, purple; Fig. 2, top). Finally, the effect of Sex on vaccine effectiveness for each of the 3 post-vaccination time periods was calculated as 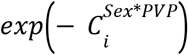.

### Ethics committee approval

The study protocol was approved by the ethics committee of Maccabi Healthcare Services, Tel-Aviv, Israel. The IRB includes an exempt from informed consent.

## Data Availability

According to the Israel Ministry of Health regulations, individual-level data cannot be shared openly. Specific requests for remote access to de-identified data should be referred to Maccabitech, Maccabi Healthcare Services Institute for Research & Innovation.

## Supplementary Figure Captions

**Supplementary Figure 1:**
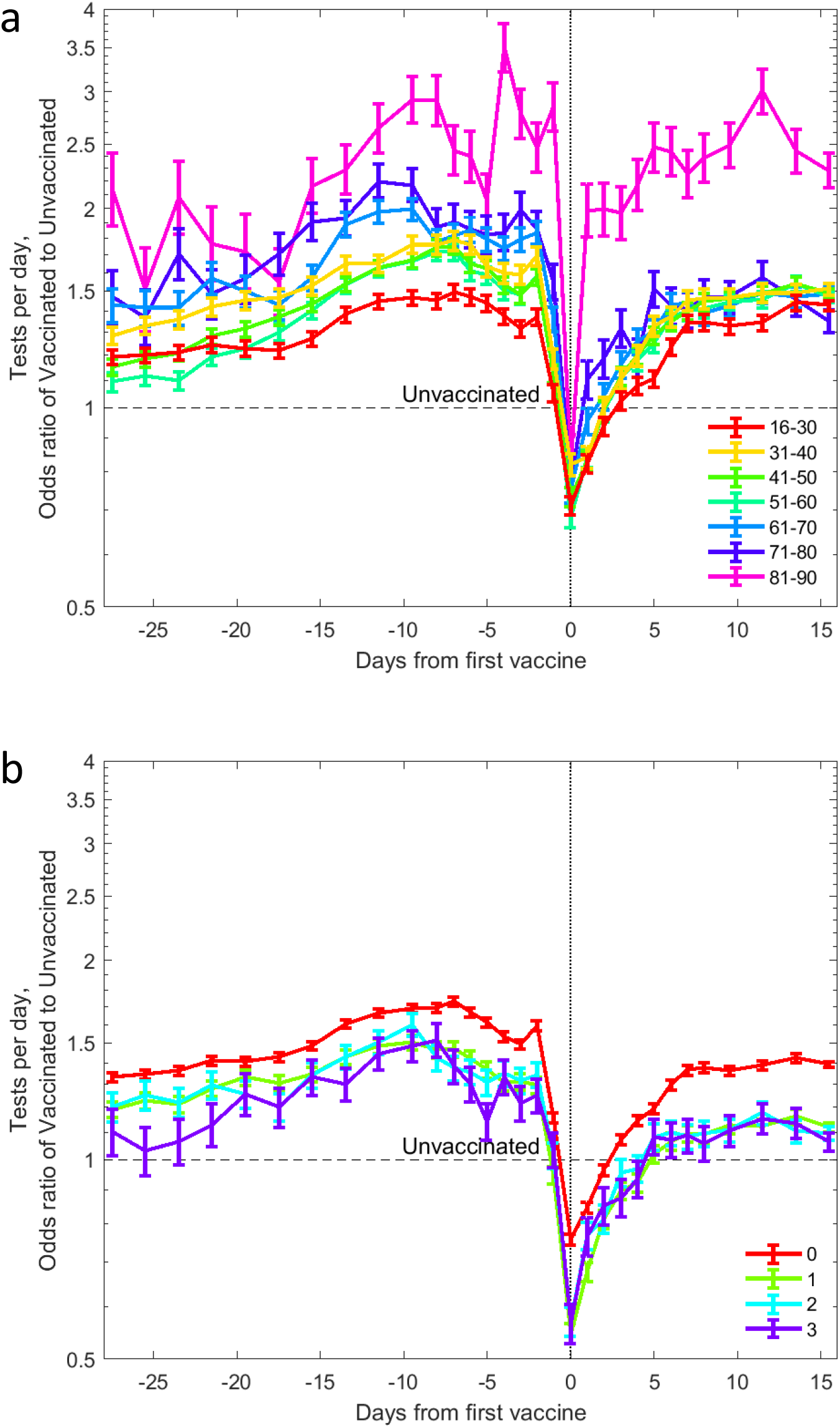
Odds ratio for testing relative to unvaccinated reference as a function of time prior and following vaccination. Results of logistic regression models for tests per day for different age groups (a) and different number of comorbidities (b). Odds ratio of tests, realive to unvaccinated, are plotted for times pre- (negative) and post- (positive) vaccination.

**Supplementary Fig. 2:**
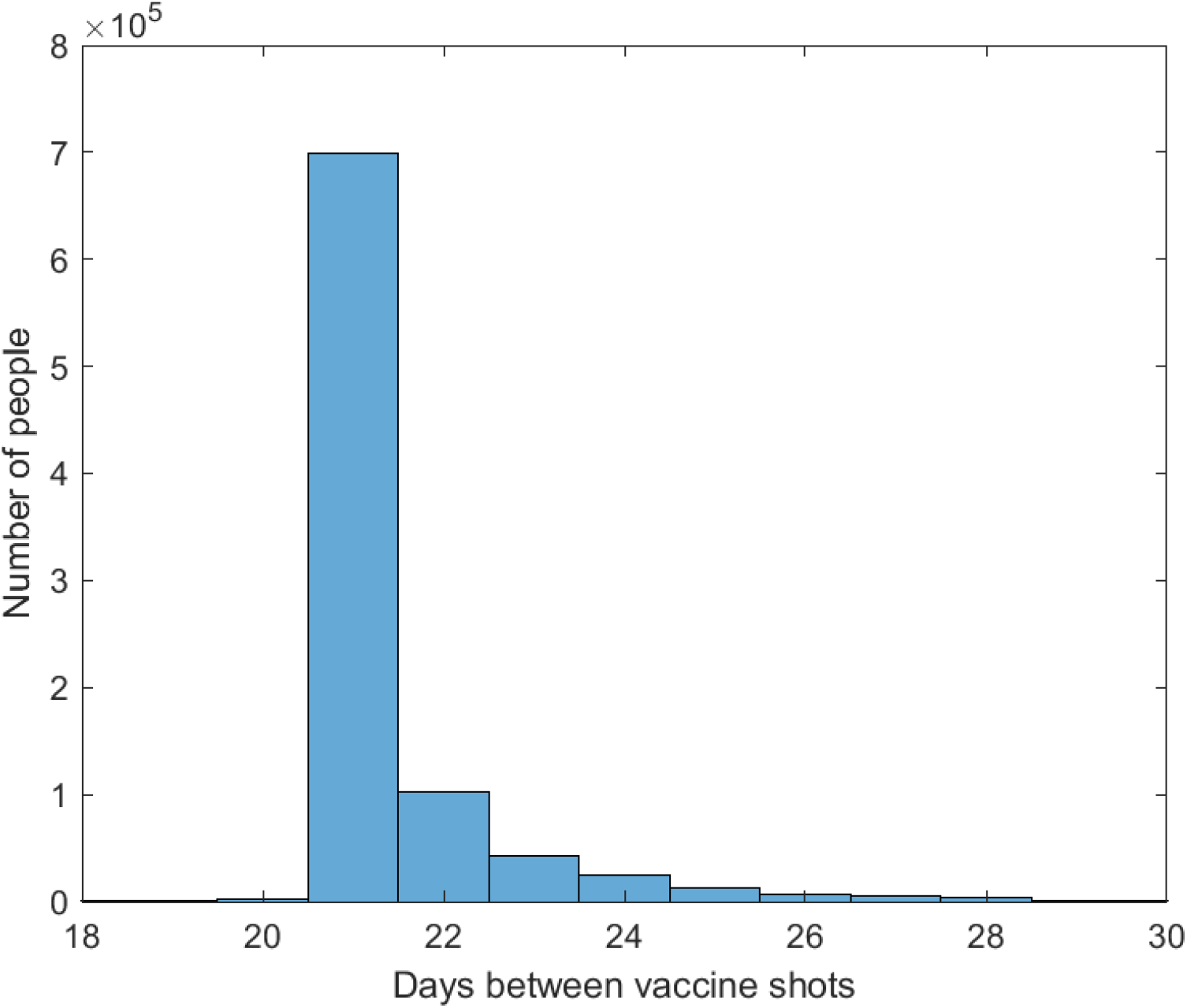
High adherence in second dose timing. Number of patients receiving the second dose as a function of days since first inoculation. In total, 98.2% of patients administered with the first vaccine dose were inoculated with a second dose by day 30.

**Supplementary Fig. 3:**
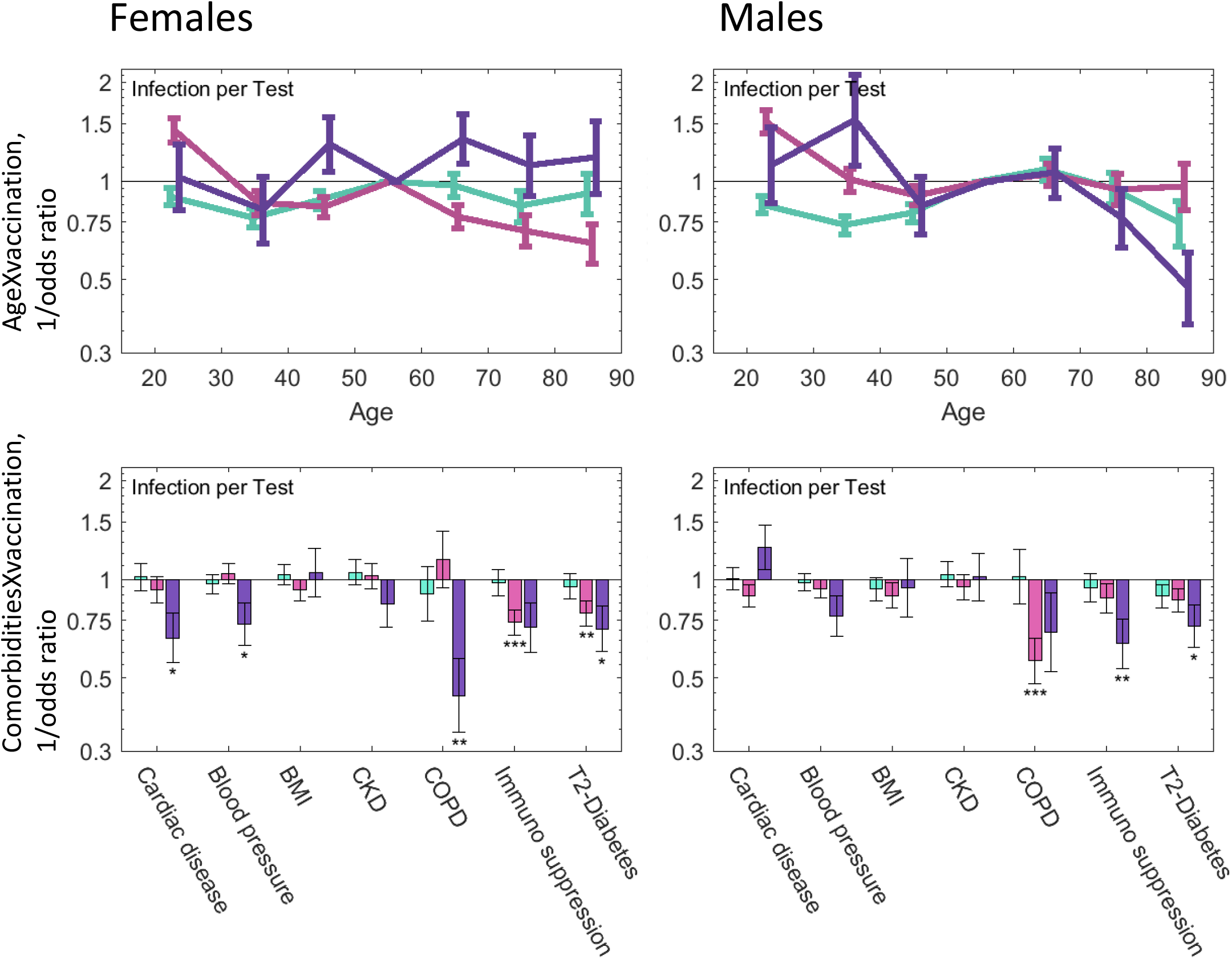
Odds ratio of infection for the interaction of age groups and comorbidities with vaccination, separated by sex. Odds ratio for per-test infections, for age (top) and comorbidities (bottom), for both females (left) and males (right), for three post-vaccination time periods: 1-11 (cyan), 12-28 (magenta) and 29-50 days (purple). Values higher than 1 (horizontal line) indicate greater vaccine effectiveness, while values lower than 1 indicate lower effectiveness. Error bars indicate one standard error. * p<0.05, ** p<0.01, *** p<0.001.

## Notes

### Competing Interest Statement

The authors have declared no competing interest.

### Funding Statement

This work was supported by the ISRAEL SCIENCE FOUNDATION (grant No. 3633/19) within the KillCorona-Curbing Coronavirus Research Program.

### Summary of Updates

This version includes the results of additional analysis of the same dataset including interactions between sex and comobidities and sex specific effectiveness by age. Additionally, figure colors have been improved.

